# A rapid, high-sensitivity SARS-CoV-2 nucleocapsid immunoassay to aid diagnosis of acute COVID-19 at the point of care

**DOI:** 10.1101/2020.12.11.20238410

**Authors:** Paul K. Drain, Madhavi Ampajwala, Christopher Chappel, Andre B. Gvozden, Melanie Hoppers, Melody Wang, Robert Rosen, Stephen Young, Edward Zissman, Michalina Montano

## Abstract

**Background:** The LumiraDx SARS-CoV-2 antigen test, which uses a high-sensitivity, microfluidic immunoassay to detect the nucleocapsid protein of SARS-CoV-2, was evaluated for diagnosing acute COVID-19 in adults and children across point-of-care settings.

**Methods:** Two paired anterior nasal swabs or two paired nasopharyngeal swabs were collected from each participant. Swabs were tested by the LumiraDx SARS-CoV-2 antigen test and compared with real-time PCR (rt-PCR; Roche cobas 6800 platform). Positive- and negative predictive values and likelihood ratios were calculated. Results stratified based on gender, age, duration of symptoms, and rt-PCR cycle threshold.

**Results:** Out of the 512 participants, aged 0-90 years, of this prospective validation study, 414 (81%) were symptomatic for COVID-19 and 123 (24%) swabs were positive for SARS-CoV-2 based on rt-PCR testing. Compared with rt-PCR, the 12-minute swab test had 97.6% sensitivity and 96.6% specificity within 12 days of symptom onset, representing the period of infectivity. All (100%) samples detected within 33 rt-PCR cycles were also identified using the antigen test. Results were consistent across age and gender. Despite being performed by minimally trained healthcare workers, the user error rate of the test system was 1%.

**Conclusion:** The rapid high-sensitivity assay using nasopharyngeal or anterior nasal sampling may offer significant improvements for diagnosing acute SARS-CoV-2 infection in clinic- and community-based settings.

**Summary:** A 12-minute nasal swab test detects 97.6% of COVID-19 infections, compared to gold standard real-time PCR testing, up to 12 days following symptom onset using a microfluidic immunoassay for SARS-CoV-2 nucleocapsid protein.

## Introduction

Severe acute respiratory syndrome coronavirus2 (SARS-CoV-2) emerged in Wuhan, China in December 2019 and rapidly spread across the world [1,2]. As of November 2020, the World Health Organization had reported over 55 million confirmed cases and over 1.3 million deaths [3]. However, due to challenges with employing diagnostic testing, the reported numbers may significantly underestimate the global burden of SARS-CoV-2 [4].

Until now, laboratory testing has focused on detecting sequences of the SARS-CoV-2 RNA genome from a nasopharyngeal swab, but this approach has several major limitations. First, laboratory testing remains laborious and expensive, which may limit access for underserved and vulnerable populations. Second, a slow turnaround time for receiving laboratory-based results may delay a person’s ability to self-isolate and prevent transmission [5,6]. Third, existing diagnostic labs have had limited supply of molecular reagents, while low- and middle-income countries have limited capacity to scale-up nucleic acid testing, to meet the needs of their communities [7]. Thus, there remains an urgent need for rapid diagnostic testing of acute SARS-CoV-2 infection in clinic- and community-based settings [8].

SARS-CoV-2 has four major structural proteins, including nucleocapsid (N), spike (S), membrane (M), and small envelope (E). The N protein, which is highly phosphorylated, interacts with SARS-CoV-2 RNA and makes up the viral core and nucleocapsid [9]. The N protein is a highly conserved target of the SARS-CoV-2 nucleocapsid, and therefore allows for reliable detection and quantitation. Our objective was to evaluate a rapid, high-sensitivity immunoassay to detect the N protein of SARS-CoV-2 for use in clinic- and community-based acute COVID-19 testing programs [10].

The LumiraDx SARS-CoV-2 antigen test runs on a portable, wall outlet or battery-powered multi-assay point-of-care instrument (Fig. 1A) [11]. The assay reagents are dry single-use, disposable, microfluidic test strips that contain specific antibodies to form an immunoassay complex that uses a fluorescent latex signal to detect the N protein of SARS-CoV-2 in a test sample. Nasal swab samples are extracted using the extraction buffer and a transferred vial dropper that delivers 20 μL onto a test strip. The SARS-CoV-2 Ag test takes 12 minutes to deliver a positive or negative result after the sample test strip has been inserted into the instrument. The instrument platform has a touch-screen interface (Fig. 1B), and connects to a cloud server for uploading test data into electronic medical records.

**Fig 1.**
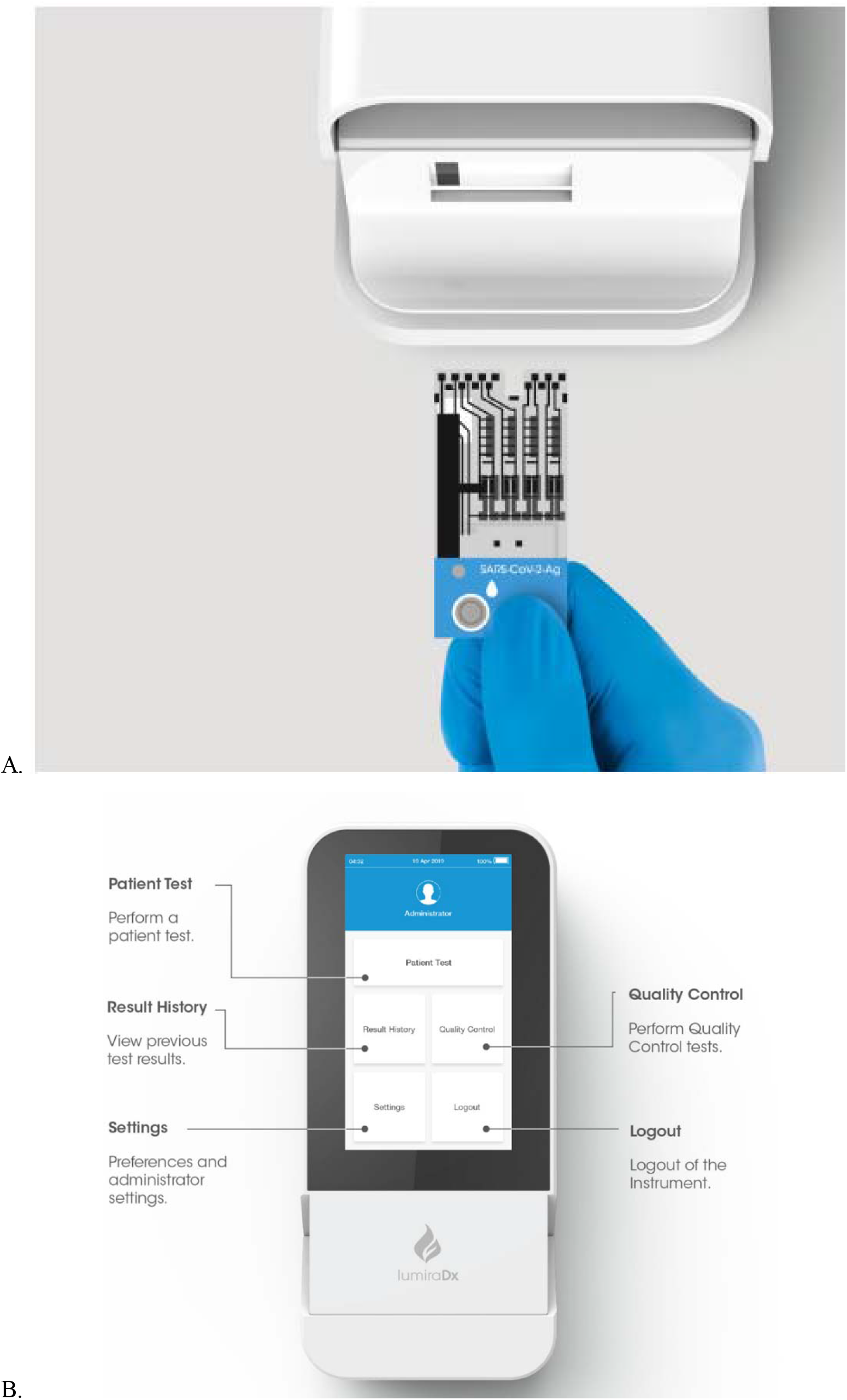
LumiraDx SARS-CoV-2 antigen assay. Test strip (**A**) and instrument home screen (**B**)

The assay limit of detection (LoD), which was established using reciprocal dilutions of gamma-irradiated SARS-CoV-2, isolate USA WA1/2020, was estimated as 32 TCID50/mL (*11*). The assay cross-reactivity was evaluated by testing a panel of microorganisms that may have high prevalence for people being tested for SARS-CoV-2. Sixteen viruses, 11 bacteria, and two fungi were tested in the absence or presence of heat inactivated SARS-CoV-2 at 3-fold LoD, and no interference was detected with pooled human nasal wash or among this panel of microorganisms [11]. The assay was also evaluated by potential endogenous and exogenous interfering substances, including anti-viral medications and over-the-counter cold remedies. Among the 22 substances tested, there was no interference with assay results [11].

A prospective cohort study was conducted to evaluate the LumiraDx SARS-CoV-2 antigen assay among children and adults who presented for COVID-19 testing.

## Materials and Methods

### Clinical validation study

A prospective validation study of the LumiraDx SARS-CoV-2 Ag Test was conducted among children and adults who presented for COVID-19 testing. The study was conducted at nine sites across the United States and United Kingdom, and including seven sites in which minimally trained operators collected and tested specimens. The clinical study received ethical approval from WCG Institutional Review Board and all participants provided informed consent (NCT 04557046 clinicaltrials.gov). Nasal swab samples were provided by a commercial supplier (MRN Diagnostics, Florida, USA), and also collected from an at-risk population (LumiraDx Stirling, UK), under approved protocols and informed consent. After collecting clinical data, two paired anterior nasal swabs (Copan FLOQ swabs) or two paired nasopharyngeal swabs were collected from each participant. The two swabs were collected simultaneously from both anterior nares, and both nostrils were swabbed using each of the two swabs. Each swab entered one nostril as the first-pass swab, before the swabs were switched to enter the opposite nostril as the second-pass swab.

Then, one swab was placed into 0.7 ml of a proprietary extraction buffer for LumiraDx SARS-CoV-2 antigen test, and the other swab was placed into 3 ml of viral transport media (VTM). Swabs in VTM were tested by rt-PCR using the SARS-CoV-2 assay using a Roche cobas 6800 platform (Roche Molecular Diagnostics, Indianapolis, IN, USA). All buffer specimens for anterior nasal swabs were tested at the clinical site, and then frozen within one hour of nasal swab collection. They were subsequently retested in a blinded manner. All buffer specimens for nasopharyngeal swabs were tested fresh at the clinical site within one hour of collection.

### Sample size and statistical analysis

Based on an anticipated diagnostic sensitivity of 95%, we targeted a sample size to ensure at least 80 positive specimens for the anterior nasal swabs to achieve a 95% confidence interval from 87-98%. For nasopharyngeal swab specimens, we targeted 40 samples to meet the minimum EUA requirement by the FDA. We evaluated diagnostic performance and stratified results by gender, age, duration of symptoms, and rt-PCR cycle threshold. We also calculated positive predictive values, negative predictive values, positive likelihood ratios, and negative likelihood ratios. We calculated 95% confidence intervals using a Wilson 2-sided analysis [12].

### Operator usability study

Eight health care workers completed a 12-question Intended Use Operator Questionnaire, which evaluated various metrics of test usability and safety. Each question was assessed on a 5-point Likert scale, ranging from 1=strongly disagree to 5=strongly agree (Supplementary Fig. S1). The responses of eight test operators, who performed participant tests using the LumiraDx Diagnostic Platform and SARS-CoV-2 Ag test, are summarized in supplementary Fig. S1.

## Results

### Participant characteristics

Among 512 participants, ages ranged from 0-90 years and 287 (56%) were female (Supplementary Table S1). Four hundred and fourteen (81%) participants experienced symptoms consistent for COVID-19 with an average duration of four days at the time of testing. Based on the Roche cobas rt-PCR testing results, 83 nasal swabs were positive for SARS-CoV-2, and 40 nasopharyngeal swabs were positive for SARS-CoV-2, giving an overall estimated prevalence of 24% for COVID-19 in this cohort.

### SARS-CoV-2 antigen assay clinical validation

Overall, the LumiraDx SARS-CoV-2 antigen assay had a sensitivity of 97.6% (95% CI 91.6-99.3%) and specificity of 96.6% (95% CI 92.7-98.4%) up to 12 days post symptom onset for nasal swab samples, and sensitivity of 97.5% (95% CI 87.1-99.6%) and specificity of 97.7% (95% CI 94.7 – 99.0%) for nasopharyngeal swab specimens (Table 1). When restricted to people testing within 10 days of symptom onset, which likely correlates to a period of viral culturability *(13)*, the diagnostic sensitivity was 98.7% (95% CI 93.0-99.8%) for nasal swabs (Supplementary Table S2). There were no appreciable differences when results were stratified by age or gender. The SARS-CoV-2 antigen assay was highly sensitive up to and including a Ct value of 33 cycles (Table 1). As expected, the rt-PCR Ct values increased with more days since the onset of symptoms (Fig. 2). In addition, Ct values >30 cycles were not uncommon shortly after symptom onset, which highlights the need for a high sensitivity test to identify individuals with a low viral load. Among participants who had an rt-PCR Ct value ≤33 cycles, the SARS-CoV-2 antigen assay was 100% sensitive for both nasal and nasopharyngeal swab specimen types.

**Table 1.** Diagnostic performance of the LumiraDx SARS-CoV-2 antigen assay nasal and nasopharyngeal swabs compared to RT-PCR for clinical testing. CI=Confidence Interval;

|  | Anterior Nasal Swab<br>n=257 |  |  | Nasopharyngeal Swab<br>n=255 |  |  |
| --- | --- | --- | --- | --- | --- | --- |
|  | Sensitivity<br>% (CI) | Specificity<br>% (CI) | LR+ | Sensitivity<br>% (CI) | Specificity<br>% (CI) | LR+ |
| Total Cohort | 97.6 (91.6-99.3) | 96.6 (92.7-98.4) | 28.3 | 97.5 (87.1-99.0) | 97.7 (94.7-99.0) | 41.9 |
| Sex |  |  |  |  |  |  |
| Females | 96.2 (87.2-99.0) | 96.6 (90.6-98.8) | 28.5 | 100 (86.7-100) | 99.2 (95.4-99.9) | 120.0 |
| Males | 100 (88.6-100) | 96.5 (90.1-98.8) | 28.3 | 93.3 (70.2-98.8) | 95.8 (89.7-98.4) | 22.2 |
| Age |  |  |  |  |  |  |
| ≤60 years | 97.4 (91.1-99.3) | 96.4 (92.3-98.3) | 26.8 | 97.4 (86.8-99.5) | 97.4 (94.1-98.9) | 38.0 |
| >60 years | 100 (56.6-100) | 100 (70.1-100) | N/A | 100 (20.7-100) | 100 (83.9-100) | N/A |
| Rt-PCR Ct Threshold |  |  |  |  |  |  |
| ≤33 cycles | 100 (94.0-100) | N/A | N/A | 100 (91.0-100) | N/A | N/A |
| >33 cycles | 60 (23.1-88.2) | N/A | N/A | 0.0 (0.0-79.3) | N/A | N/A |

LR=Likelihood Ratio; N/A=Not available

**Fig 2.**
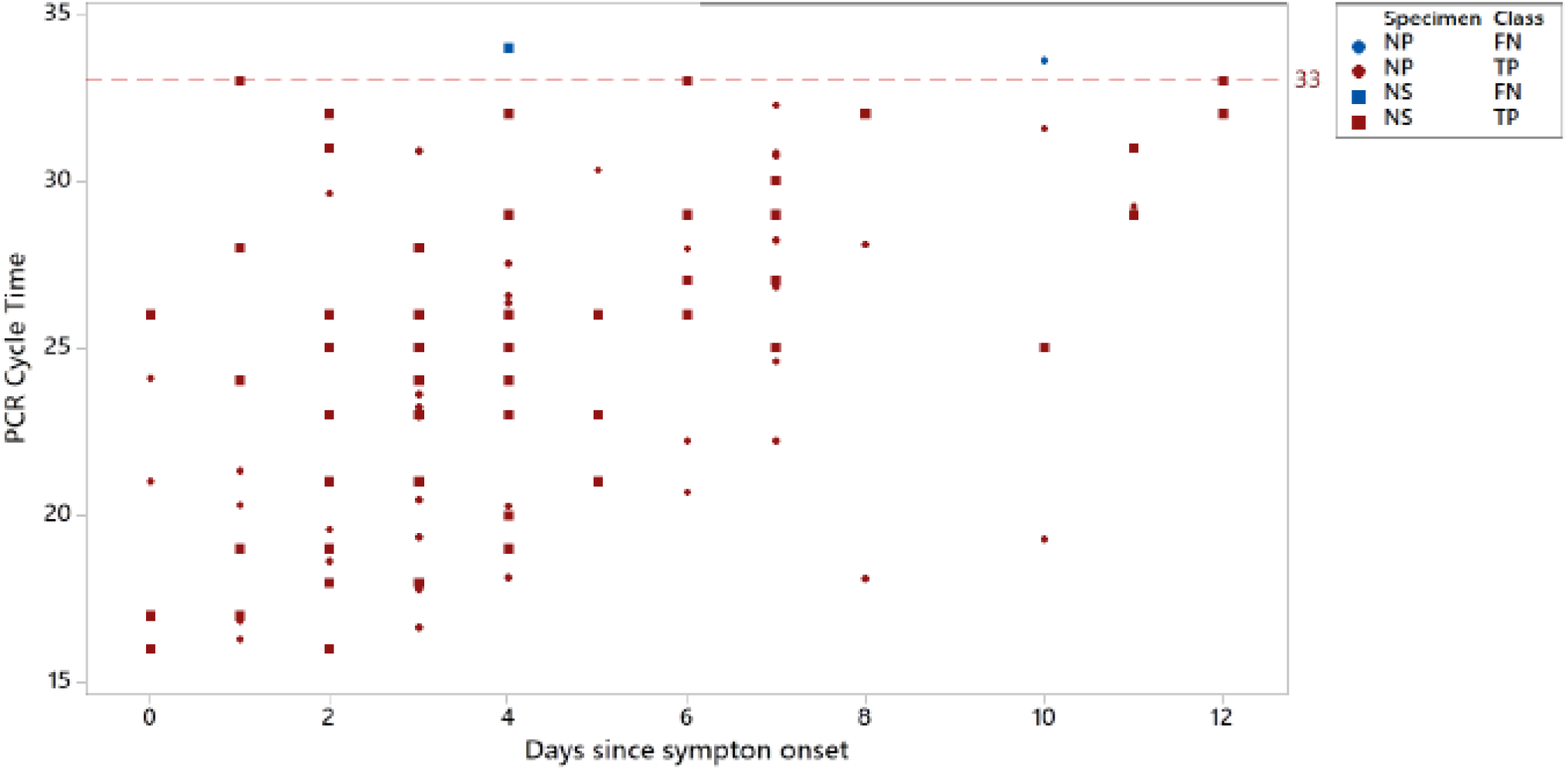
Roche cobas SARS-CoV-2 rt-PCR cycle threshold versus days since symptom onset. True positive (TP) and false negative (FN) results using the LumiraDx SARS-CoV-2 antigen test with nasal (NS) or nasopharyngeal (NP) swab specimens. Red circles and squares indicate participants positive by LumiraDx SARS-CoV-2 antigen test. Blue circles and squares indicate participants negative by LumiraDx SARS-CoV-2 antigen test. Dotted line represents Ct 33, the limit of detection for the LumiraDx SARS-CoV-2 antigen test.

### SARS-CoV-2 antigen assay usability

During the prospective study, tests were completed by minimally trained healthcare workers who performed field-based testing at various sites including a drive thru test center, community testing hub, as well as pediatric and family medicine clinics. Usability was assessed by monitoring user errors and obtaining feedback via questionnaire. The user error rate of the test system was recorded at 1%. Overall, positive responses were obtained for ease of use of the test system, easy to follow instructions and simple to interpret results (Supplementary Fig. S1).

## Discussion

At the outset of the study, Emergency Use Authorization (EUA) for the test was sought from the US Food and Drug Administration (FDA) to cover the following: strip and sample stability, strip and sample freeze-thaw, limit of detection (LoD), analytical specificity, microbial and substance interference, high-dose hook, and point-of-care use [10]. Based on the data submitted to the FDA, the LumiraDx SARS-CoV-2 antigen assay with nasal swab received EUA on August 18, 2020 (*14*).

Several other rapid SARS-CoV-2 antigen tests have received EUA from the FDA for use in near-patient settings. Two have limited diagnostic sensitivity (85-88%) when used within five days since symptom onset [14]. A second assay demonstrated high diagnostic sensitivity, but was only authorized for the first seven days since symptom onset [14]. In contrast, this study demonstrated that the LumiraDx SARS-CoV-2 antigen assay had high diagnostic sensitivity, particularly with rt-PCT Ct values of ≤33 cycles, as measured by the Roche cobas SARS-CoV-2 rt-PCR assay, through the first 12 days from the onset of COVID-related symptoms.

Recent studies have identified the first 10 days since symptom onset as the likely window of infectivity for the SARS-CoV-2 virus [13,15,16]. Several other studies have related infectivity to a low rt-PCR Ct value, which correlates to a high viral load, and/or the ability to culture SARS-CoV-2 virus [17-21]. A study of hospitalized COVID-19 patients showed the ability to culture SARS-CoV-2 diminished from day 10 to day 12 since symptom onset [13]. In another study of hospitalized patients, successful isolation of culturable virus correlated with rt-PCT Ct values of ≤33 cycles, while those above this level were considered to be non-infectious [19]. rt-PCR testing has also been shown to remain positive many days past the window for culturing viable SARS-CoV-2 virus, which suggest that rt-PCR testing may be generating some positive results from people with remnant viral RNA who do not have contagious viral particles [20].

In summary, the LumiraDx SARS-CoV-2 antigen assay demonstrated high sensitivity when used to diagnose COVID-19 at the clinical point of care. The rapid assay was highly sensitive for people with rt-PCR Ct values of ≤33 cycles within a period of 12 days since the onset of COVID-19 symptoms. These performance characteristics may correlate well with reported infective SARS-CoV-2 viral load and window of infectivity. The assay achieved a lower LoD and higher diagnostic sensitivity than other POC tests and provided fast results (in under 12 minutes), in a convenient, easy to use point-of-care test format, with capacity to transfer data to electronic health records and surveillance systems.

In conclusion, there is an urgent need to improve access to point-of-care testing for SARS-CoV-2 during the COVID-19 pandemic. This test demonstrated high sensitivity over a wide range of rt-PCR Ct values up to and including a Ct value of 33 cycles, and over a 12-day infectivity window, making this platform highly suitable for SARS-CoV-2 testing and COVID-19 surveillance programs. This rapid assay with high sensitivity and anterior nasal sampling offers significant advantages for identification and management of SARS-CoV-2 infection, particularly in clinic- and community-based settings.

## Data Availability

Data and materials availability: The study dataset may be provided upon
request.

## Acknowledgments

We thank the children, women and men who participated in this study, the clinical sites for sharing their space, and our research staff and nurses who conducted the study. The content is solely the responsibility of the authors.

## Author contributions

All authors contributed to the interpretation of the data, critically revised the manuscript for important intellectual content, approved the final version to be published, and agree to be accountable for all aspects of the work.

