## Supplemental material for "A rapid, high-sensitivity SARS-CoV-2 nucleocapsid immunoassay to aid diagnosis of acute COVID-19 at the point of care"

Supplementary Materials:

**Supplemental Table 1.** Cohort for the validation study of LumiraDx SARS-CoV-2 antigen assay.

|  | **Anterior Nasal swab**  **n= 257** | **Nasopharyngeal Swab**  **n= 255** |
| --- | --- | --- |
|  | **Mean (SD) or N (%)** | **Mean (SD) or N (%)** |
| Age (years): *mean (SD)* | 34 (+/-15.7) | 33.2 +/- 19.4 |
| Age categorical |  |  |
| ≤ 5 years | 13 (5.1%) | 22 (8.6%) |
| 6 to 21 years | 29 (11.3%) | 59 (23.1%) |
| 22 to 59 years | 200 (77.8%) | 150 (58.8%) |
| ≥ 60 years | 15 (5.8%) | 24 (9.4%) |
| Female Gender | 142 (55.3%) | 145 (56.9%) |
| Clinical Signs and Symptoms |  |  |
| Number with COVID signs/symptoms | 159 (61.9%) | 255 (100%) |
| Days since symptom onset - mean (SD) | 4.0 (2.9) | 3.5 (2.5) |
| POC and laboratory testing |  |  |
| Positive LumiraDx SARS-CoV-2 antigen test | 87 (33.9%) | 44 (17.3%) |
| Positive Roche cobas SARS-CoV-2 rt-PCR | 83 (32.3%) | 40 (15.7%) |

SD=standard deviation

**Supplemental Table 2.** Diagnostic sensitivity of the LumiraDx SARS-CoV-2 antigen assay by days since symptom onset.

|  | ***Anterior Nasal Swab*** | | ***Nasopharyngeal swab*** | |
| --- | --- | --- | --- | --- |
| **Days Since  Symptom Onset** | ***#TP/***  ***(#TP+#FN)*** | **Sensitivity**  **% (95% CI)** | ***#TP/***  ***(#TP+#FN)*** | **Sensitivity**  **% (95% CI)** |
| 0 | 6/6 | 100 (61.0-100) | 2/2 | 100 (34.2-100) |
| ≤ 1 | 12/12 | 100 (75.8-100) | 6/6 | 100 (61.0-100) |
| ≤ 2 | 28/28 | 100 (87.9-100) | 9/9 | 100 (70.1-100) |
| ≤ 3 | 37/37 | 100 (90.6-100) | 17/17 | 100 (81.6-100) |
| ≤ 4 | 54/55 | 98.2 (90.4-99.7) | 22/22 | 100 (85.1-100) |
| ≤ 5 | 60/61 | 98.4 (91.3-99.7) | 23/23 | 100 (85.7-100) |
| ≤ 6 | 66/67 | 98.5 (92.0-99.7) | 26/26 | 100 (87.1-100) |
| ≤ 7 | 72/73 | 98.6 (92.6-99.8) | 34/34 | 100 (89.8-100) |
| ≤ 8 | 74/75 | 98.7 (92.8-99.8) | 36/36 | 100 (90.4-100) |
| ≤ 9 | 74/75 | 98.7 (92.8-99.8) | 36/36 | 100 (90.4-100) |
| ≤ 10 | 76/77 | 98.7 (93.0-99.8) | 38/39 | 97.4 (86.8-99.5) |
| ≤ 11 | 79/80 | 98.8 (93.3-99.8) | 39/40 | 97.5 (87.1-99.6) |
| ≤ 12 | 81/83 | 97.6 (91.6-99.3) | 39/40 | 97.5 (87.1-99.6) |

CI=Confidence Interval; FN=false negative; TP=true positive

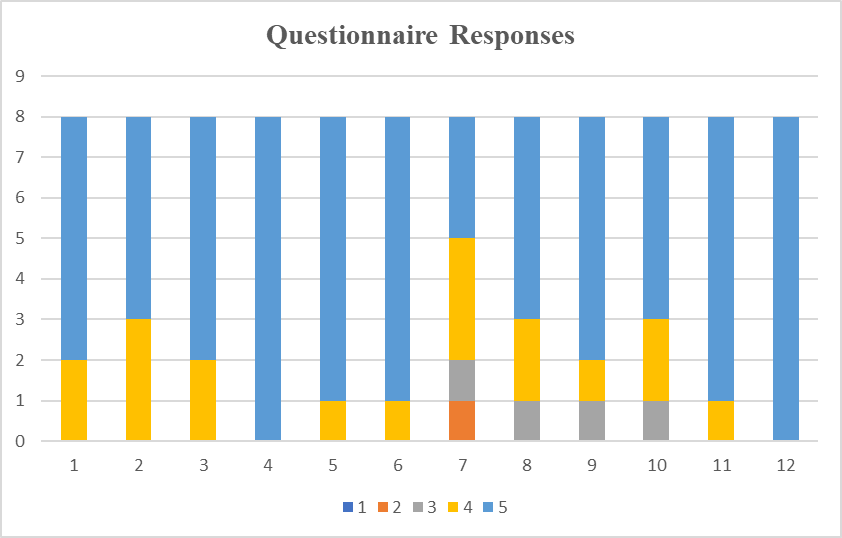

**Supplemental Fig 1.** Usability responses from minimally trained test operators. Each question was assessed on a 5-point Likert scale, ranging from 1=strongly disagree to 5=strongly agree. The survey questions were the following: (1) The Quick Reference Instructions were easy to follow; (2) The Package Insert Instructions were easy to follow; (3) The User Manual was easy to follow; (4) The on-screen test step instructions were easy to follow; (5) The test was easy to perform; (6) The results displayed on-screen at test completion were easy to interpret; (7) When running a patient test for the first time, I was able to run the test without help from anyone else; (8) Inserting the Test Strip into the LumiraDx Instrument was easy; (9) Extracting the sample was easy; (10) Adding the sample to the Test Strip was easy; (11) Removing the Test Strip was easy; (12) No safety issues were encountered.
